# Association Between Lateral Posterior Tibial Slope and Anterior Cruciate Ligament Injury: Cross-sectional Study in a Philippine Tertiary Hospital

**DOI:** 10.1101/2025.07.30.25332473

**Authors:** Terence Aaron L. Burgo, Brian Andrich L. Pollo

**Affiliations:** Department of Radiology, Manila Doctors Hospital; Biomedical Innovations Research for Translational Health Science (BIRTHS) Laboratory, Department of Biochemistry and Molecular Biology, College of Medicine, University of the Philippines Manila

**Keywords:** Anterior Cruciate Ligament Injuries, Magnetic Resonance Imaging, Risk Factors, Knee Injuries, Philippines

## Abstract

**Introduction:** Anterior cruciate ligament (ACL) injuries are common in orthopedic and radiologic practice, often caused by abrupt directional changes or trauma. Recent studies suggest that anatomical factors, such as posterior tibial slope, particularly the lateral component (LPTS), influence ACL injury risk by increasing anterior tibial translation.

**Objective:** To correlate the LPTS with ACL injury incidence at Manila Doctors Hospital and compare LPTS values between injured and non-injured groups.

**Methodology:** A retrospective cross-sectional study reviewed MRI scans of patients who underwent knee MRI from January to December 2018. LPTS was measured using the circle method on images acquired via a Siemens 1.5 Tesla MRI scanner. Cases were included if MRI was interpreted by Philippine College of Radiology-accredited radiologists.

**Results:** Of 242 participants, 225 met inclusion criteria: 179 with ACL injury (79.6%) and 46 without (20.4%). The injured group had a mean LPTS of 10.13° (SD: 2.40), significantly higher than the non-injured group mean of 7.23° (SD: 1.31) (p < 0.0001). Patients with a large LPTS were 37.69 times more likely to have an ACL injury (OR = 37.69). Among injured patients, 140 had a large LPTS versus 4 in the non-injured group.

**Conclusion:** A steep lateral posterior tibial slope is significantly associated with ACL injury. Identifying this anatomical risk factor may guide preventive strategies or early intervention in high-risk individuals.

## Introduction

In both orthopedic and radiologic practice, anterior cruciate ligament (ACL) injuries represent the most frequently encountered ligamentous injury of the knee.^1^ The ACL may be stretched, partially, or completely torn, often as a result of sudden directional changes, rapid deceleration, awkward landings, or direct trauma to the knee. Diagnosis is typically made through physical examination and confirmed by magnetic resonance imaging (MRI), which offers a non-invasive and reliable method of assessing soft tissue integrity and joint morphology. ^3^

ACL injuries carry serious functional and long-term health consequences. ^2^ Time away from sports or occupational activity can range from 9 to 12 months, with unpredictable outcomes. For instance, in a study involving professional football players, only 65% returned to top-level competition three years after an ACL tear. Furthermore, such injuries significantly increase the risk of developing early-onset knee osteoarthritis, contributing to chronic morbidity and healthcare burden.

In response to this clinical significance, many studies have explored potential anatomical risk factors that predispose individuals to ACL injuries. Among these are morphological features such as the shape and dimensions of the intercondylar notch, condylar width, quadriceps angle, α and β angles, and particularly, the posterior tibial slope (PTS). ^1,3^ The PTS refers to the angle formed between a line perpendicular to the tibial shaft and the posterior inclination of the tibial plateau. It is usually measured separately on the lateral and medial aspects of the tibial plateau (LPTS and MPTS, respectively). A steeper posterior tibial slope has been associated with increased anterior tibial translation under load and elevated strain on the ACL, especially during weight-bearing or pivoting maneuvers. ^4^ Despite these observations, current literature presents conflicting results, with some studies emphasizing the LPTS as a major risk factor, while others do not find significant associations. ^3,4^

The inconsistencies may, in part, be due to population differences. Anatomical parameters such as PTS have been shown to vary by ethnicity and sex. For example, a study by Salvatore et al. reported that Black subjects had higher tibial plateau angles compared to White individuals, with similar trends observed in Asian populations, who exhibited significantly steeper slopes than other groups. ^6^ However, even within the same sex or ethnic subgroup, findings on the relationship between tibial slope and ACL injury risk have been inconsistent. ^5^

Given these uncertainties and the lack of data in the Philippine context, this study aims to investigate the relationship between a steep lateral posterior tibial slope and the incidence of ACL injury among patients undergoing knee MRI at Manila Doctors Hospital. Specifically, it seeks to (1) identify patients with ACL injuries, (2) determine the period prevalence of ACL injury within the institution, (3) examine the correlation between a steep LPTS and ACL injury, and (4) compare LPTS values between injured and non-injured patients. Establishing this relationship may contribute to better screening of high-risk individuals and inform future preventive or therapeutic strategies tailored to the local population.

## Methods

This retrospective cross-sectional study reviewed the MRI results of all patients of Manila Doctors Hospital who underwent knee MRI from January 2018 to December 2018 as interpreted by Radiologists accredited by the Philippine College Radiology and Fellows of the CT-MRI society of the Philippines utilizing Siemens 1.5 Tesla MRI scanner.

### Ethics

This study was submitted and approved by the Technical and Ethical Review Board of the institution for review. As a retrospective study and data analysis was performed anonymously, this study was exempted from the required informed consent of patients. Formal permission to collect and access data banks was acquired from the data privacy officer to safeguard the confidentiality and integrity of patients’ data. Patients were assigned by random numbers to ensure anonymity.

### Study population

Case records of all patients who underwent knee MRI studies in the hospital from January 1, 2018 to December 31, 2018.

### Sample size estimation

Total number of all cases that were documented in 2018, as stated above were included in this study. The sample size was computed with 95 % confidence level, an acceptable margin of error at 5 % an estimated prevalence of ACL injury in large lateral posterior tibial slope of 56.8 % based on Shen, L., et al. The computation for sample size suggested a need for at least 148 patients.

### Data collection

MRI reports and images of all patients who underwent knee MRI from January 1 to December 31, 2018 were retrieved from the NOVARAD RIS and PACS system of the Department of Radiology of the Manila Doctors Hospital. Images were selected based on inclusion and exclusion criteria and the lateral posterior slope was measured by the primary investigator using the method exactly as suggested by Lipps et al and Hudek et al, under the supervision of a board-certified Radiologist and Fellow of the CTMRI society of the Philippines. The data was tabulated using Microsoft Excel-based data abstraction tool.

### Inclusion Criteria

All patients who underwent knee MRI from January 2018 to December 2018 at the Manila Doctors Hospital.

### Exclusion Criteria

1. Patients with other ligament injuries (MCL, PCL, LCL).
2. History of previous ACL injury and repair.
3. Presence of tibial or femoral fracture.
4. Patients with co-existing arthropathy and skeletal congenital abnormalities.

### Measurement of Lateral Posterior Tibial Slope

There are 2 measurements for assessing the PTS, the lateral and medial PTS. This study dealt with the LPTS measurement. The value of the LPTS has a more significant effect on knee stability than the MPTS as studies claimed.

Tibial slope is commonly defined as the angle between a line fit to the posterior-inferior surface of the tibial plateau and a tibial anatomic reference line.^7^

Sagittal-plane MRIs were obtained using Siemens 1.5 Tesla MRI scanner utilizing knee coil and neutral knee position. The sagittal plane images were used for measurements (field of view, 140 cm; slice thickness/gap: 4 mm/0.3 mm; matrix: 256×256 ; scan time, 3 minutes 45 seconds). All knee geometry measurements were performed using the Novarad DICOM software package. The most repeatable MRI method for measuring lateral tibial slope uses the “circle method” (Fig 1), which involves drawing two circles within the proximal tibial in the sagittal plane. The proximal circle was fit within the proximal, anterior, and posterior cortical borders. The center of the distal circle was positioned on the perimeter of the proximal circle, and was fit within the anterior and posterior cortices. A line connecting the centers of these two circles defined the tibial proximal anatomical axis (TPAA).^7^

**Figure 1.**
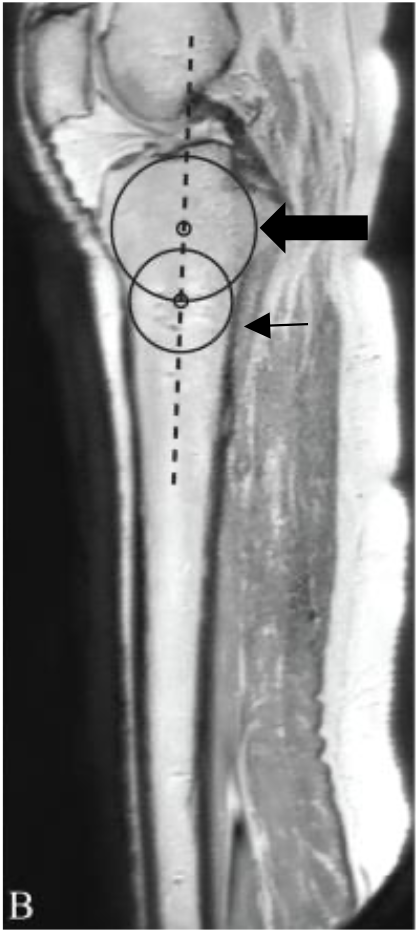
Circle method. Proximal circle (fat arrow) was fit within the proximal, anterior, and posterior cortical borders. The center of the distal circle (thin arrow) was positioned on the perimeter of the proximal circle, and was fit within the anterior and posterior cortices. A line (dashed black line) connecting the centers of these two circles defined the TPAA.

All lateral tibial slope measurements were defined as the angle between this lateral tibial plateau line and a line perpendicular to the TPAA as determined by using the “circle method” (Fig 2). The normal lateral posterior tibial plateau is 5.0º± 3.6º.^8^

**Figure 2.**
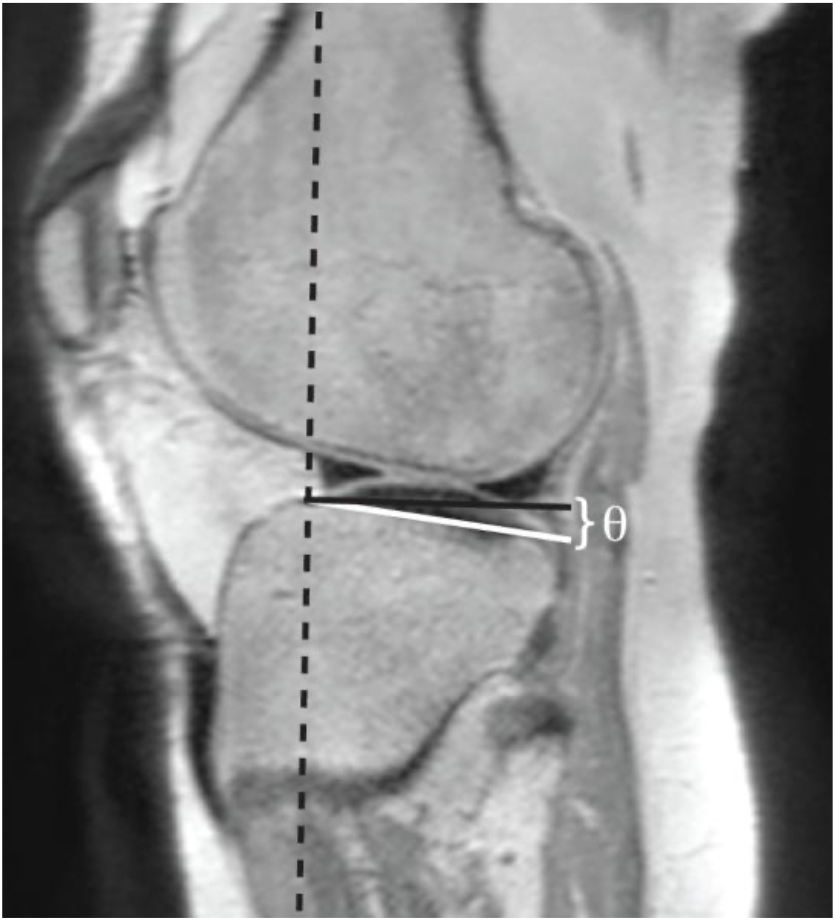
Lateral tibial slope is the angulation (θ) between a line fit to the subchondral bone line (white line) and a line perpendicular to the tibial proximal anatomic axis (black line).

### Statistical Analysis

The data analysis consisted of descriptive and inferential statistics and simple linear regression modelling as used by Shen, Lei, et al.^3^ Descriptive statistics were used to describe the demographic characteristics and MRI findings of the cases. Mean in millimeters (±SD) were calculated for all continuous variables, while proportions were estimated for all categorical variables.

The relationships between ACL injury as the dependent variable and steep lateral posterior tibial slope as the potential risk factor as independent variable were assessed by simple linear regression analysis. In that analysis, the dependent variable was defined as ACL status (0 = normal; 1 = injury). The statistical significance of the coefficients in the regression equation were determined with the Wald test. Odds ratios (*OR*s) and their respective 95% confidence intervals (*CI*s) were also estimated. All statistical analyses were performed using SPSS 24.0 (SPSS Inc., Chicago, IL, USA), and a value of *P* < 0.05 was considered statistically significant. Single factor analysis was utilized to compare the LPTS of the ACL-injured and non-injured groups

## Results

Out of the 242 knee MRI scans reviewed, 225 cases met the inclusion criteria (Table 1). Among these, a substantial majority (79.56%, n = 179) exhibited anterior cruciate ligament (ACL) injuries, while only 20.44% (n = 46) showed no signs of ACL injury. This distribution highlights the predominance of ACL injuries in the study population from Manila Doctors Hospital during the year 2018.

**Table 1.** The number of participants with injured and non-injured ACL included in the study who underwent knee MRI at Manila Doctors Hospital from January 2018 to December 2018.

| Knee MRI | Injured ACL | Non-Injured ACL | Total |
| --- | --- | --- | --- |
| Number | 179 (79.56%) | 46 (20.44%) | 225 (100%) |

Analysis of the lateral posterior tibial slope (LPTS) revealed a marked difference between the two groups (Table 2). Individuals with ACL injuries had a significantly steeper mean slope (10.13°) compared to those without ACL injury (7.23°). The variability within each group, as indicated by the standard deviation, was greater in the injured group (SD = 2.40) than in the non-injured group (SD = 1.31), suggesting a wider range of slope values among those with ACL injuries. Confidence intervals for both groups were narrow and did not overlap, further supporting the distinction between them. Statistical comparison of the mean LPTS between groups yielded a highly significant result (p < 0.0001), with a confidence interval for the difference ranging from -3.62 to -2.18 (Table 3). This indicates a robust association between a steeper tibial slope and ACL injury.

**Table 2.**
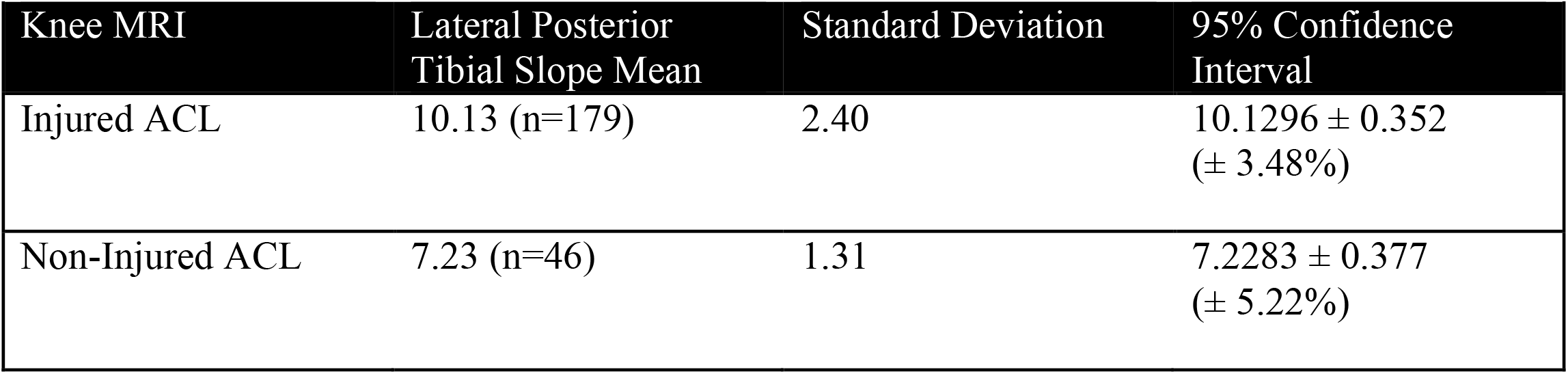
The mean, standard deviation and 95% confidence interval of participants with injured and non-injured ACL included in the study who underwent knee MRI at Manila Doctors Hospital from January 2018 to December 2018.

**Table 3.** The p-value when comparing the lateral posterior tibial slope mean of the injured and non-injured ACL groups included in the study who underwent knee MRI at Manila Doctors Hospital from January 2018 to December 2018.

| Knee MRI | Injured ACL | Non-Injured ACL | p-value | 95% Confidence Interval |
| --- | --- | --- | --- | --- |
| Lateral Posterior Tibial Slope Mean | 10.13 (n=179) | 7.23 (n=46) | p < 0.0001 | -3.6243 to -2.1757 |

When classifying the slopes as either “large” (>8.6°) or “normal,” the disparity became even more pronounced (Table 4). A majority of individuals with ACL injuries (78.2%) fell into the “large slope” category, whereas nearly all non-injured individuals (91.3%) had slopes within the normal range. The odds of having a large posterior tibial slope were 37.69 times higher in the injured group than in the non-injured group (95% CI: 12.73 to 111.58, p < 0.0001), indicating a strong association between increased posterior tibial slope and ACL injury risk.

**Table 4.** The number of the large or normal lateral posterior tibial slopes of the injured and non-injured ACL groups included in the study who underwent knee MRI at Manila Doctors Hospital from January 2018 to December 2018.

| Lateral Posterior Tibial Slope | Injured ACL | Non-Injured ACL | Odds Ratio | 95% Confidence Interval | p-value |
| --- | --- | --- | --- | --- | --- |
| Large | 140 | 4 | 37.69 | 12.7329 to 111.5781 | p < 0.0001 |
| Normal | 39 | 42 |  |  |  |

## Discussion

This study aimed to determine the association between the lateral posterior tibial slope (LPTS) and the incidence of anterior cruciate ligament (ACL) injury among patients who underwent knee MRI at Manila Doctors Hospital. Of the 242 knee MRI studies reviewed, 179 were included based on the presence of a complete tear, sprain, or partial tear of the ACL, defined radiologically as the absence of normal-appearing fibers or the presence of focal high signal or ligamentous laxity. Seventeen studies were excluded due to concomitant ligament injuries or associated bone fractures. The findings revealed that patients with a steeper LPTS were significantly more likely to have ACL injuries, with an odds ratio of 37.69 (P < 0.0001), indicating a strong association between increased posterior slope and ligament injury.

These results are consistent with prior literature that has identified the LPTS as an important anatomical risk factor for ACL tears. Prior studies have shown that increased LPTS correlates with greater anterior tibial translation during dynamic loading, thereby increasing mechanical strain on the ACL ^3^. This biomechanical relationship explains why individuals with steeper tibial slopes are more susceptible to ACL injuries, especially during sports or trauma involving pivoting, deceleration, or jumping. The ACL serves to resist anterior translation of the tibia, and when the posterior slope is exaggerated, the tibia is more likely to move forward under load, potentially overwhelming the stabilizing capacity of the ligament.

The mean LPTS for ACL-injured patients in this study (10.13° ± 0.35) was slightly lower than values reported in some previous studies but remained elevated relative to typical reference values. This variation could be attributed to anatomical differences influenced by ethnicity, sex, or age. For example, differences in tibial slope among South Indian populations have been observed, ^10^ and degenerative changes and sex have been found to significantly influence PTS measurements. ^11^ Similarly, women have been found to tend to have higher medial and lateral slopes compared to men, with racial differences also noted. ^4^ Despite these variations, there are currently no established normative LPTS values for the Filipino population, highlighting the importance of localized research.

A small subset of non-injured patients in this study also demonstrated elevated LPTS values, while some ACL-injured patients had normal slopes. This finding is consistent with the multifactorial nature of ACL injury, where anatomical risk factors such as LPTS interact with other intrinsic and extrinsic variables, including neuromuscular control, activity level, joint laxity, and external forces. Although these inconsistencies exist, the significantly elevated odds of ACL injury in patients with steep LPTS support its role as a contributing factor. The generalizability of the findings here may be limited to similar populations. Anatomical norms vary significantly across ethnic groups, and prior studies have demonstrated that tibial slope values differ among Asian, Black, and White populations. Therefore, these findings are most applicable to urban Filipino or Southeast Asian populations and may not fully extrapolate to other demographic groups without further validation

This study had several limitations. First, there may have been selection bias related to sex distribution between the injured and control groups, which was not controlled for. Some evidence suggests that men with higher LPTS may be more susceptible to ACL injuries than women with similar slope values. Second, the analysis did not adjust for other biomechanical, neuromuscular, or behavioral factors that may influence ACL injury risk. Third, the study did not explore whether greater degrees of LPTS were associated with more severe ACL damage. Lastly, measurement was based on retrospective MRI data from a single institution and scanner, which may limit reproducibility.

Future studies should aim to establish normative LPTS values specific to the Filipino population and investigate potential cutoff points that may predict ACL injury. Prospective, multi-center studies with sex-stratified analysis and biomechanical modeling would further elucidate the role of LPTS in knee injury. Clinicians and radiologists should consider including tibial slope measurements in routine knee MRI evaluations, particularly in at-risk individuals, to inform preventive strategies and surgical planning.

## Conclusion

This study demonstrates a significant association between a steep lateral posterior tibial slope and the incidence of anterior cruciate ligament injury among patients at Manila Doctors Hospital. Individuals with higher LPTS values were substantially more likely to sustain ACL injuries, supporting existing biomechanical theories and prior research. While LPTS alone is not the sole determinant of injury, it is an anatomical risk factor that may help identify individuals at greater risk. Given the lack of normative data in the Filipino population, further research is warranted to establish population-specific thresholds and to explore the interplay between LPTS and other intrinsic and extrinsic risk factors. Incorporating LPTS assessment into routine knee MRI interpretation may aid in early risk stratification, preventive counseling, and individualized management strategies.

## Data Availability

All data produced are available online at https://doi.org/10.7910/DVN/XQGZ2I.

## Ethics committee approval

This report was conducted in accordance with institutional ethical standards of Manila Doctors Hospital. Its institutional review board waived ethical committee approval.

## Authors contribution statement

Terence Burgo was responsible for Investigation, Resources, Supervision, Writing – Original Draft, and Writing – Review & Editing. Brian Pollo contributed to contributed to Conceptualization, Data Curation, Literature Review, Writing – Original Draft, and Writing – Review & Editing. Both authors approved the final version of the manuscript.

## Competing interests

The authors declare no competing interests.

## Funding

No funding was received.

## Data availability statement

All photographs and radiographs pertaining to this study are available in the Harvard Dataverse repository at https://doi.org/10.7910/DVN/XQGZ2I.

## Notes

### Competing Interest Statement

The authors have declared no competing interest.

### Funding Statement

This study did not receive any funding

### Author Declarations

Institutional Review Board of Manila Doctors Hospital waived ethical approval for this work

## References

1. Chiu SSH. The anterior tibial translocation sign. Radiology. 2006;239(3):914–915. doi:10.1148/RADIOL.2393040273,

2. Rekik RN, Tabben M, Eirale C, et al. ACL injury incidence, severity and patterns in professional male soccer players in a Middle Eastern league. BMJ Open Sport Exerc Med. 2018;4(1). doi:10.1136/BMJSEM-2018-000461,

3. Shen L, Jin ZG, Dong QR, Li LB. Anatomical Risk Factors of Anterior Cruciate Ligament Injury. Chin Med J (Engl). 2018;131(24):2960. doi:10.4103/0366-6999.247207

4. Weinberg DS, Williamson DFK, Gebhart JJ, Knapik DM, Voos JE. Differences in Medial and Lateral Posterior Tibial Slope. American Journal of Sports Medicine. 2017;45(1):106–113. doi:10.1177/0363546516662449,

5. Wordeman SC, Quatman CE, Kaeding CC, Hewett TE. In vivo evidence for tibial plateau slope as a risk factor for anterior cruciate ligament injury: A systematic review and meta-analysis. American Journal of Sports Medicine. 2012;40(7):1673–1681. doi:10.1177/0363546512442307,

6. Bisicchia S, Scordo GM, Prins J, Tudisco C. Do ethnicity and gender influence posterior tibial slope? Journal of Orthopaedics and Traumatology. 2017;18(4):319–324. doi:10.1007/S10195-017-0443-1,

7. Lipps DB, Wilson AM, Ashton-Miller JA, Wojtys EM. Evaluation of different methods for measuring lateral tibial slope using magnetic resonance imaging. American Journal of Sports Medicine. 2012;40(12):2731–2736. doi:10.1177/0363546512461749,

8. Hudek R, Schmutz S, Regenfelder F, Fuchs B, Koch PP. Novel Measurement Technique of the Tibial Slope on Conventional MRI. Clin Orthop Relat Res. 2009;467(8):2066. doi:10.1007/S11999-009-0711-3

9. Sims K. Musculoskeletal MRI. J Can Chiropr Assoc. 2010;54(2):134. Accessed July 24, 2025. https://pmc.ncbi.nlm.nih.gov/articles/PMC2875914/

10. Nekkanti S, Patted P, Nair L, Chandru V, Shashank G. The variation of the posterior tibial slope in South Indians: A hospital-based study of 290 cases. Nigerian Journal of Orthopaedics and Trauma. 2018;17(1):17. doi:10.4103/NJOT.NJOT_4_18

11. Pangaud C, Laumonerie P, Dagneaux L, et al. Measurement of the Posterior Tibial Slope Depends on Ethnicity, Sex, and Lower Limb Alignment: A Computed Tomography Analysis of 378 Healthy Participants. Orthop J Sports Med. 2020;8(1). doi:10.1177/2325967119895258,

